# Co designing a novel game for young people who have experienced adverse childhood experiences: Protocol

**DOI:** 10.1101/2023.12.27.23300570

**Authors:** Isabelle Butcher, Mina Fazel, Paul McCrone, Tanya Krzywinska, Anton Belinskiy, Lindsay Smith, Minhua Ma, Gabriela Pavarini, Harsimran Sansoy, Anna Mankee-Williams, Ben Teasdale, Kamaldeep Bhui

## Abstract

**Background:** Adverse childhood experiences (ACEs) are associated with poorer mental health outcomes later in life. Research to date has focused on utilising qualitative and quantitative measures to understand possible mechanisms, and interventions for those who may have experienced ACEs.

**Objectives:** This study aims to co-design a novel digital game, in collaboration with youth and adult stakeholders, to engage 12-25-year olds in learning about the impact of ACEs and how to protect and promote mental health following ACE exposure. Additionally, this project aims to evaluate the feasibility and acceptability of such an intervention.

**Methods:** This study will adopt Experience Based Co- Design methods with adolescents aged between 12 and 24 living in England, and will involve adult stakeholders working in the public sector. We will then evaluate the feasibility and acceptability of the co-designed game on young people, and test its cost-effectiveness.

**Conclusion:** This project will enable an insight into the processes of co-designing a youth-informed public mental health game-based intervention and feasibility and acceptability of a serious game approach for adolescents aged between 12-24 years of age who may have experienced ACEs.

**Strengths and limitations of this study:**

- Adverse childhood experiences (ACEs) are associated with poorer mental health outcomes later in life. Research to date has primarily focused on quantitative and qualitative data to explore possible resources for young people. This study describes a novel digital game study for young people who may have experienced adversities.
- The data on adverse childhood experiences has primarily on focused on urban and rural areas; this study seeks to recruit young people from rural, urban and coastal regions across England
- This study is restricted to England, and whilst it is hoped the findings will be applicable to the U.K and more globally, the authors acknowledge the limitations of working with young people only in England
- This study is one of a few studies that has sought to involve young people through the co design of a resource for young people who may have experienced adversities from initial conception of the idea to implementation of the game.

## Introduction

Adverse Childhood Experiences (ACEs) are identified as key experiences in the life of a child or adolescent from which change-both positive and negative-might take place for that individual (1, 2). ACEs refer to three types of difficult experiences, those related to abuse (verbal, sexual, or physical); neglect; and household challenges (parental separation or incarceration; substance misuse, domestic violence, or family mental illness). Other important adversities that are not officially categorised as ACEs but are significant include victimisation (bullying, peer rejection, racism), the consequences of living in socioeconomic deprivation (poverty, living in unsafe environments) and multiple traumatic losses, early death of caregiver, harsh experiences in care, and poor academic performance. ACEs, by their very nature, cluster in socio-economically deprived areas, and can have broad impacts including on mental health outcomes across the lifespan, and of note, contribute to a reduced life expectancy by up to two decades(3). Of adolescents exposed to multiple ACEs, up to 3 in 4 will develop mental health problems by the age of 18, including major depression, conduct disorder, post-traumatic stress disorder and alcohol and substance misuse in addition to behaviours of self-harm and suicide attempts). Furthermore, health care costs are substantially increased for those experiencing ACEs.

Young people who have experienced multiple ACEs can present with complex trauma-associated symptoms including avoidance and emotional dysregulation(4), which can make emotional processing and recall as well as engagement harder. For some, seeking support and the prospect of what might then happen can be overwhelming leading to avoidance of therapeutic engagement and interventions, in fear that they will not be able to manage their emotions or memories if explored or that the person they are talking to will not be able to help them. In this context, it is important to design and implement new ways to assist those who are finding it difficult to access or engage with existing services and interventions to access support-either through changing how support can be accessed or developing new interventions to address the fears and concerns that can become the drivers of avoidance. Engagement can therefore be a particularly complex component of interventions for those who have experienced ACEs and is therefore an important area to focus on-especially for younger populations. not just in the initial stages of a treatment but throughout the process.

Game-based interventions offer a novel and potentially promising approach in youth mental health, given the heightened interests of young people in these forms of interactive technology. Serious games can enable adolescents to interact through play in order to cultivate skills, knowledge, and judgement relevant to game’s focus, including the promotion of physical activity, social-emotional development and different types of psychological and physical states. Serious games have been used and found to be helpful for the treatment of a range of mental health difficulties, including ADHD and depression. Game-based interventions have been used in therapeutic work for trauma with adults, and there is some research showing benefit for young people experiencing ACEs.

We report on a study developing new interventions and methods of engagement for those who have experienced ACEs through creative arts, community based participatory research as well as utilising advances in digital technologies. The study co-designs and tests a digital game to specifically benefit young people who are least likely to engage with traditional services, such as those who have experienced ACEs across diverse identities and place contexts, including neurodivergent groups.

### Research questions

1. Is a co-designed, youth-informed public mental health game-based intervention acceptable and feasible for young people who have experienced ACEs?
2. What are the benefits of the serious game-based interventions for the mental health of adolescents who have experienced ACEs?
3. What are the mechanisms by which the serious game influences the experience and response to ACEs?
4. Do game based interventions represent potential value for money?
5. What ethical considerations do serious games raise when used with young people?
6. Is the designed game accessible and acceptable to a broad range of young people from diverse identities living in rural, urban and coastal places?

### Research Objectives

1. To engage young people of diverse identities from diverse contexts in creative ‘sprints’ to establish the broad parameters of the structure, options, and ethically pertinent features of a novel digital game.
2. To engage young people in a process of co-design to shape the content (images, narratives, scenarios) of the game which will likely include a range of potential scenarios, some leading to positive and helpful outcomes, alongside others that require different types of support.
3. To evaluate the acceptability, efficacy and engagement with the game by assessing take up; level of sustained use; attention to specific content, feedback on helpful and unhelpful elements, using standardised outcome measures at baseline, after a period of game play, and at a 3 month follow up.
4. To conduct a process evaluation to assess whether the game is empowering, offers control, is engaging, and can be personalised as well as explore mechanisms of avoidance, emotional resilience, the intersectional influences by gender, sexuality, ethnicity, place, and neurodiversity.
5. To assess the cost effectiveness of this game intervention.

### Aims

1. To co-design a novel digital game, in collaboration with youth and adult stakeholders, to engage 12-25-year olds in learning about the impact of ACEs and how to protect and promote mental health following ACE exposure.
2. To evaluate the acceptability and feasibility of a newly developed serious game on 12-24 years old who have experience of ACEs.

### Ethics

This study has received ethical approval from the UK National Health Service Health Research Authority (23/WM/0105 June 2023).

## Methods

### Patient and public involvement

Patients and/or the public have been involved in the design, and continue to be involved in the design, conduct, reporting or dissemination plans of this research.

### Recruitment

Young people (age 12 – 16 and 16–24) from diverse identity subgroups will be recruited through relevant organisations; LGBTQIA+ community, ethnic minority, neurodivergent, marginalised groups such as refugees, gypsy and traveller community, and by place of residence (rural/coastal/urban; Cornwall, Kent, London, Leeds, Central England Central England (Berkshire/Buckinghamshire/Gloucestershire/Oxfordshire). We seek to recruit young people in different contexts of education (included/excluded), some in contact with formal health services, and some with care experiences. This is a purposive sample seeking representation from the identified groups and not a representative sample of any one area or their general population. The purpose is to bring the insights from these groups and those at the intersections of these groups, anticipating that they might benefit from the developed game. The aspiration is to include people with a diverse range of experience of mental health and illness, as well as those previously exposed to multiple adverse events.

## Sample size

### Serious Game co-design (technological and exploring potential functions)/ Game Development Procedure

The games design team will meet with young people on a six weekly cycle and explore design options. Eight workshops will be conducted in total. This will provide time to factor in design recommendations, and to produce prototypes including personalisation options, new creative content, functionality, ethical dimensions and attractiveness to the diverse groups included. Each stage will include documentation of young people’s feedback; design decisions following on from the co-design meetings will be documented. The usability, acceptability, and feasibility of addressing all the aims and objectives will be documented to inform the experience-based co-design group. These sessions will be audio recorded and photographs and videos may be taken.

Co-design will take place with a group of approximately 15 young people (aged 12-24 years). Co-design sessions with the youth co-design team will be held through the period of game development. We plan to host approximately 12 sessions – 8 dedicated to general design and 4 to ethical dimensions. Depending on young people’s availability and the need for further feedback on the same topic, some of the sessions will be conducted twice. The sessions will be held virtually via Teams primarily and sometimes in person; each will last between one and two hours. Sessions will be video or audio recorded.

Learnings and summaries of data from previous work packages (1 and 2) within the wider Attune project will be fed in as part of the work of the work package 4 (WP4) and will be led by researchers working on WP4 across the ATTUNE programme. The game development team will meet weekly with clinicians to gain a more in-depth understanding of young people’s experience of ACEs. Meetings with young people also took place earlier once the game development team were recruited, Figure 1 outlines this process.

### Serious Game: Experience Based Co-design

Approximately 15 young people will be recruited from across the project representing different stakeholder groups via ATTUNE partner organisations. An expression of interest flyer will be used. They will represent the same diverse groups as specified above. Furthermore, we will also recruit 5-10 adult stakeholders (teachers-school and university, parents, employers, social workers, mental health professionals, primary care professionals). We will ask for recommendations across the ATTUNE team and reach relevant stakeholders directly via email. The approach taken will be of experience-based co-design, using material from earlier work packages (experience data from creative arts workshops and statistical analyses, synthesised into easy ready materials for presentation to the Experience Based Co-Design (EBCD) group. The group will hear the summary material and will have access to more detailed materials (anonymised) if required.

We propose to hold one group which will be held in a hybrid format to enable participation for those who are unable to travel and will plan to hold a single group with representation from each region. We will aim for one EBCD group of 2-3 sessions with 10 adults and 15 YP. Preferably in person or online or possibly hybrid depending on preference. There will be adequate breaks, refreshments, and ‘chill out’ spaces. It is important to note that these workshops may be conducted over a longer intensive accelerated process over two days. The first meeting will aim to be face to face, with the possibility of later meetings being held online. The preference is for in person meeting(s), as often communication is better, power-relationships can be more easily explored and managed, and participants can interact with examples of the creative outputs and data analyses, summarised. Co-design processes need to be responsible to participants with whom power is shared, therefore, despite this intended plan, we will retain flexibility to respond to the needs of young people and stakeholders who may prefer fewer or more sessions, and in smaller or larger groups. We are also working on the principles of information power, that is we need sufficient data to achieve our purpose, when saturation might be achieved, rather than continue if additional workshops are not adding new knowledge or perspectives.

The sessions will be audio and video recorded. All participants will be provided with an electronic information sheet and consent processes, and young people will be also asked to complete our baseline questionnaires from which we can determine levels of health problems and need for support during the process. This information will be securely held on the University of Oxford servers, and reviewed weekly, to determine if any protocols need to be actioned.

**Figure 1.**
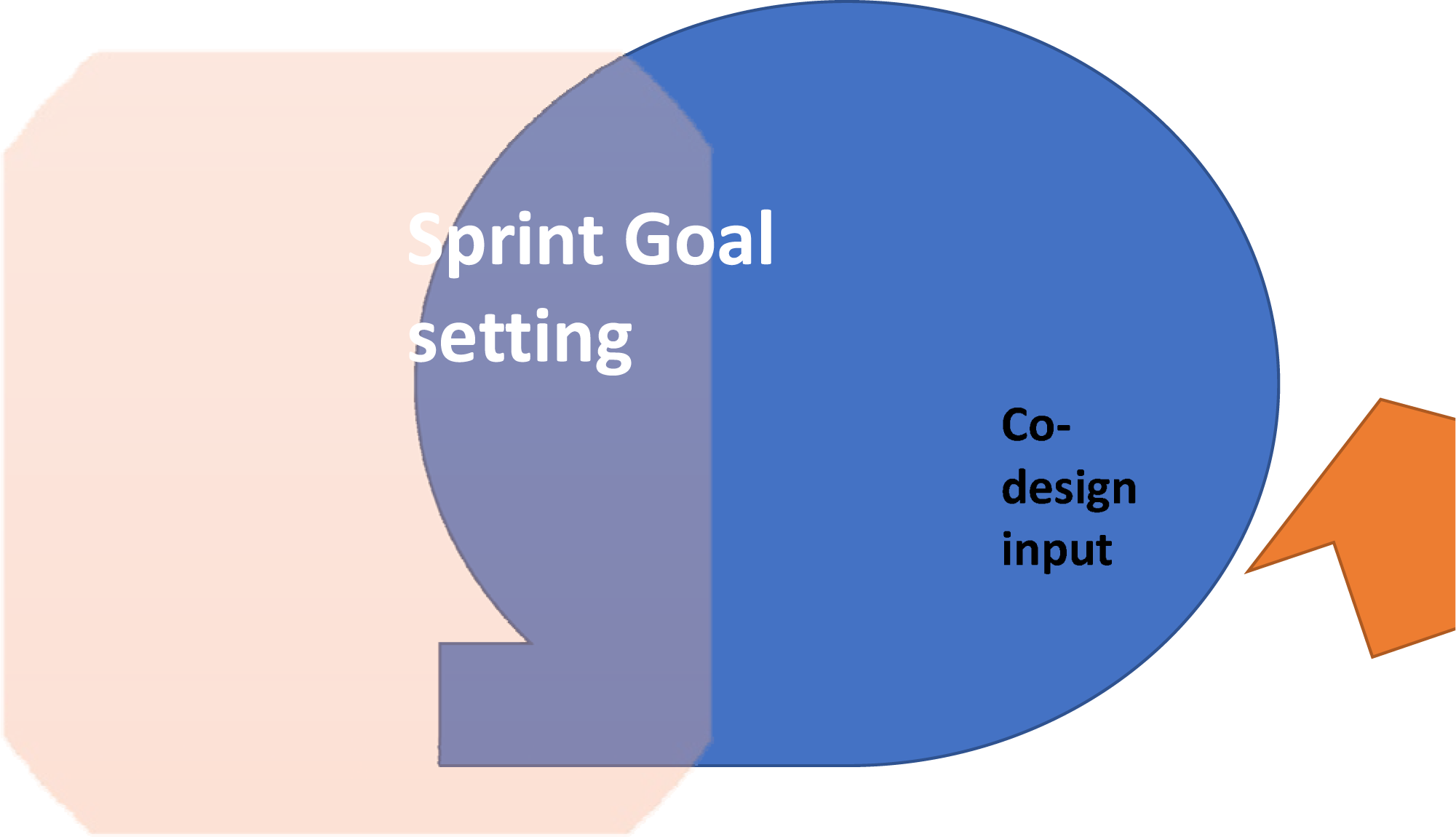
The Agile method of game development is designed to enable iteration and therefore facilitates deep co-design. *It works with sprints, i.e.; periods of defined development*.

### Acceptability and Feasibility Evaluation

The acceptability and feasibility study will broadly follow MRC guidance on the evaluation of complex interventions (5, 6).Approximately 40 young will be recruited via ATTUNE partner organisations operating in Cornwall/Kent/Oxford/Leeds. The participants will not have been involved in any other parts of the ATTUNE project to form an independent and unbiased sample for evaluation.

An expression of interest flyer will be used. Information sessions about the study will be held at each organisation (either in person or online) and/or key workers within the organisation will approach participants and introduce them to the project. Young people will be recruited from each of the ATTUNE study sites across the country to then test the developed intervention. They will first complete the online consent process, supported locally by the organisation through which they are recruited. We will seek carer/parental consents where appropriate, and support entry by pen and paper forms if required.

After consent (or assent with parental consent), young people will be individually screened by a researcher, where they will complete a brief demographic questionnaire and an ACEs measure. Those aged 12-24 years who report having experienced 3 or more ACEs will be eligible to be selected into the sample of 40 young people testing the intervention. We anticipate recruiting up to 60 young people to take account of drop-out and also to ensure balance in demographic characteristics (age, gender, ethnicity, sexuality, neurodiversity, place, levels of severity). Recruitment will include community organisations and NHS mental health team wait lists.

The participants will be aged 12-24 (dependent on game content and guidance on age groups suitable content). Data will be collected on who the game is appropriate for, and whether it meets the needs of all these groups though audio recordings/photos/videos. For those screened into the study an additional measure of ACEs will be used to understand the presence/absence, frequency/severity and any other adverse life events not assessed by the screening tool. Evaluation will include numbers recruited and retained, levels of usage, and completion of outcome measures.

Outcome measures: questionnaires will be completed at baseline, immediately post-intervention, and at 3-month follow up. We will collect quantitative data and assess pre-post differences of mean scores and distributions on validated measures of wellbeing (Warwick Edinburgh Mental Wellbeing Scale-7 item suitable for YP (aged 15-21), post traumatic symptoms (Revised Impact of Events Scale-13 items covering reexperiencing, avoidance, hyperarousal), anxiety (GAD-7), depression (PHQ-9), self-compassion (Neff Scale) and use of services and engagement with educational/social activities – these will be administered using the CSRI.

Acceptability will be assessed using Sekhon et al. (2022)’s theoretical framework(7) and checklist including *a priori* validated questions on: affective attitude, burden, ethicality, perceived effectiveness, coherence, self-efficacy, opportunity costs, general acceptability. The feasibility will be assessed by:

- success in recruitment of diverse groups (age, gender, ethnicity, sexuality, rural-coastal-urban localities),
- use of the game (frequency, duration, at least 3-5 occasions over 1-2 months; explored and finalised in the co-design phase),
- retention in the evaluation to the end,
- completion of outcome measures,
- ability of delivery mechanisms (digital and remote) to be reliable and attractive and easy to use (Likert questions designed for this study by the peer research and PPI team),
- presence of sufficient support (Likert) from intervention in-built elements as well as location of delivery (home, in school, NGO; we anticipate most will engage with the intervention at home or in a supported environment).

We will assess acceptability and feasibility of collecting quantitative outcome measures. We will assess feasibility for running a larger evaluation in the future based on the engagement and completion of questionnaires and the intervention. We will generate preliminary data on potential differences between age range, geo-social location, and types of ACEs and if this new game-based approach is feasible, acceptable, and helpful to young people of diverse identities, and the extent to which avoidance behaviours regarding accessing mental health support are changed.

Where possible the screening and all questionnaires will be conducted face-to-face with support from a trained researcher at the partner organisation, or online via Teams. In-person support will be available to those completing the questionnaires unless they have been screened as able to do this online. The young person will fill in the forms using a digital device provided (tablet or computer). When judged appropriate (e.g., for older adolescents), links to questionnaires will be sent via email and the game completed independently online.

For those completing the game independently, a brief check-in phone call with an ATTUNE researcher will be arranged during the period in which the game is tested. Specific arrangements for data collection will be decided on a case-by-case basis in consultation with the relevant partner organisation, the young person (and guardians where applicable).

As process data we will use statistics gathered computationally by the game, e.g., length of play, where a game is dropped by a user, character(s)/role(s) played, narrative choices made within the game, r, in-game signposting information accessed.

### Process Evaluation

A process evaluation will be undertaken with 20-40 young people, who participated in the evaluation, ensuring representation from those who had high, medium and low levels of engagement with the game, including those who did not engage with the intervention or dropped out (subject to consent to follow up). The information will be collected by 1:1 interview, either face to face or online via Teams, with appropriate support from i.e. key worker, youth worker, parent etc. This individual, if they wish to attend the 1:1 with the consent of the young person, will be asked to consent as a participant themselves in the interview, and may also comment on their perspectives on acceptability and feasibility of the game if they observed the young person doing it. The process evaluation(8) will explore qualitatively:

- The context of how the intervention was used (alone in their room, whilst on public transport etc….).
- The levels of support used and needed.
- The elements of the intervention used and which were helpful and which not.
- The time spent using the intervention.
- Experiences of benefit or concerns about distress and harms.
- Mechanisms of benefit or harm as explained by young people’s narrative account.
- What helped and what made it harder to use the intervention?

The data from the process analysis will be transcribed and thematically analysed using principles of Framework analysis(9). We are interested in exceptional experiences as well as norms in the analysis, and variation by different identities and places.

### Ethics decision making and personalisation

Our method adopts an embedded ethics approach to game design, whereby ethically relevant issues are assessed and discussed with stakeholders throughout the whole process, with the game adjusted accordingly to ensure responsible innovation. This includes issues of representativeness, accessibility, emotional safety, responsible motivation, and responsible mental health/ACEs content, among others. Informed consent will be obtained from all youth (or young person’s assent along with parental consent for under 16s) and adult contributors’ participants before the study commences. All young people will be reimbursed the equivalent of the hourly living wage (10.51 pounds) per hour for their contribution.

### Analysis

The data from the process analysis will be transcribed and thematically analysed using principles of Framework analysis(9). We are interested in exceptional experiences as well as norms in the analysis, and variation by different identities and places.

### Economic evaluation

Analyses will be conducted to assess the cost-effectiveness of the intervention. The CSRI will be used to measure health and social service use at baseline and throughout the follow-up period. Time off school/college/work will also be recorded. Costs will be calculated by combining the resource use with appropriate unit cost information. The cost of the actual intervention will be estimated based on development time and ongoing refinement and support. We will investigate variations in cost across the sample using regression models and compare costs for those with different levels of change in the other measures. The relationship between costs and other outcomes will be explored using regression analyses and variations according to uptake of the intervention will be explored.

## Discussion

This study seeks to create a novel mobile-based game, designed by in relation the feedback of young people and adult stakeholders to enhance engagement with treatment for mental health problems. Furthermore, an evaluation of the impact of the game with young people who have experienced ACEs, alongside an acceptability and feasibility study will be conducted. The analysis will identify whether any particular groups may be more likely than others to benefit from the serious game approach as an intervention for their mental health. It is hoped the game will help those who have experienced a wide range of unpleasant adversities and that it will be accessible to a broad range of young people.

Evaluating the impact of this intervention and its cost effectiveness will be crucial when considering the role of game-based interventions for those working in policy and other key stakeholders. Through the setting up of young people’s advisory groups, in a range of regions and localities, including urban, rural and coastal in England this study will then disseminate findings to young people and stakeholders through established relationships and relevant formats including social media channels and existing groups and community spaces that young people attend.

## Patient consent for publication

Not applicable

## Data Availability

Data not available.

## Acknowledgements

The authors wish to thank each member of the wider ATTUNE research team who have contributed to the wider work of the project.

## Contributors

KB is the principal investigator for the ATTUNE study(ATTUNE — Department of Psychiatry (ox.ac.uk) All authors made substantial contributions to the conceptualisation and design of the study. Tk, GP, IB, MM, KB, PM drafted the first version of the manuscript, with input from other named authors. IB, TK, KB, GP, HS, MM, and MF reviewed the manuscript for intellectual content, approved the final version to be published and agreed to be accountable for all aspects of the work.

## Footnotes

i Attune (attuneproject.com)

